# Clinical features of COVID-19 patients hospitalized at the Tashkent State Medical University and risk factors for intensive care unit admission: a cross-sectional study from Uzbekistan, Central Asia

**DOI:** 10.64898/2026.08.28.26361631

**Authors:** Bakhodir Rakhimov, Jaewook Choi, Kyonghee Kim, Laziz Tuychiev, Alisher Shadmanov, Bakhromjon Mamatkulov

## Abstract

**Background:** The clinical course of coronavirus disease 2019 (COVID-19), and the ability to anticipate which patients will require intensive care, were poorly characterized in Central Asia during the first pandemic wave. We aimed to describe the clinical features of hospitalized COVID-19 patients at the Tashkent State Medical University, Uzbekistan, and to identify risk factors for intensive care unit (ICU) admission.

**Methods:** In this single-centre cross-sectional study, we reviewed the records of 2500 consecutive patients hospitalized between 11 April and 8 August 2020. Patients were grouped as asymptomatic or symptomatic, and symptomatic patients were compared by ICU versus non-ICU status. Groups were compared with chi-square or Fisher’s exact and Mann-Whitney U tests. Univariable and multivariable logistic regression identified risk factors for ICU admission.

**Results:** Of 2500 patients (median age 36 years; 60.9% male), 989 (39.6%) were asymptomatic and 1511 (60.4%) symptomatic. In total, 129 (5.2%) were admitted to the ICU and 38 (1.5%) died. ICU patients were older (median 56 vs 40.5 years) and more often had bilateral pneumonia, oxygen desaturation and cardiometabolic comorbidity. In the multivariable model (AUC 0.82), the independent predictors of ICU admission were ischemic heart disease (aOR 4.20), shortness of breath (aOR 3.22), hypertensive heart disease (aOR 2.93) and male sex (aOR 2.00).

**Conclusions:** Older age, cardiometabolic comorbidity and respiratory compromise identified patients at high ICU risk. As one of the first clinical COVID-19 descriptions from Uzbekistan, these data provide a baseline for preparedness in Central Asia.

## 1. Introduction

The disease that emerged abruptly in China in December 2019 and subsequently spread worldwide was named coronavirus disease 2019 (COVID-19), and the causative virus was recognized by the World Health Organization (WHO) as SARS-CoV-2.^1,2^

The clinical course of COVID-19 ranges from an asymptomatic form to admission to the intensive care unit (ICU).^3^ The disease proceeds mainly in a mild form, whereas approximately 20% of symptomatic patients require ICU treatment owing to complications of additional severe disease.^4,5^ Older age, male sex, symptoms such as high fever, fatigue and cough, and comorbidities including cardiovascular disease, hypertension and diabetes mellitus have been described as significant criteria for severe illness and ICU admission.^6^

One of the most urgent problems in medicine is the early detection of emergency cases, which otherwise overwhelm the health system; strengthening and expanding the capacity of primary health care can help to overcome this challenge.^7^

On 15 March 2020 the Ministry of Health of the Republic of Uzbekistan reported the first case of COVID-19, detected in a citizen returning from France. A sharp increase in incidence was noted from the beginning of June 2020, with two to six deaths per day, and the maximum number of new cases occurred in early August 2020.

Central Asia, and Uzbekistan in particular, remains substantially under-represented in the international COVID-19 literature. Although a few reports on the early situation in Uzbekistan have been published,^8^ there is an overall lack of studies in international journals that combine epidemiological, clinical, laboratory and treatment information for the first months of the pandemic in the region. Such baseline descriptions of an under-studied health system retain value beyond the first wave: they document how a resource-constrained system responded when no specific therapy was available, provide a reference point for comparison with later waves and other Central Asian countries, and inform preparedness for future respiratory-pathogen emergencies. Therefore, this study aimed to describe the clinical features of COVID-19 patients categorized into non-ICU and ICU groups in a single hospital and to identify the risk factors for ICU admission.

## 2. Methods

### 2.1. Study design and setting

This single-hospital cross-sectional study was reported in accordance with the STROBE statement for observational studies. It was performed at the Multidisciplinary Clinic of the Tashkent State Medical University, one of the largest hospitals in the Republic, with a total capacity of 1477 beds and designed for 180,000 visits per year. At the beginning of the pandemic three main hospitals in Tashkent city were re-organized to treat COVID-19 patients; the Multidisciplinary Clinic of the Tashkent State Medical University was one of them and was temporarily adapted to treat patients from April to August 2020.

### 2.2. Participants and case definitions

Selection criteria based on severity were adopted from the interim guidelines for the management of patients infected with COVID-19 (6th ed.),^9^ prepared by the Ministry of Health of the Republic of Uzbekistan with support from the WHO.^10^ Asymptomatic cases were patients with a positive SARS-CoV-2 real-time PCR result and without complaints, clinical symptoms or pathological changes on lung imaging. Patients with clinical symptoms and pathological changes were classified as symptomatic and divided into four categories: mild, moderate, severe and critical. Data from all 2500 patients hospitalized from 11 April to 8 August 2020 were collected; the last patient was discharged on 18 August 2020. Patients were divided into asymptomatic (n=989) and symptomatic (n=1511) groups; among symptomatic patients, epidemiological and clinical features were compared between non-ICU and ICU groups and risk factors for ICU admission were identified. Discharge or death was the study endpoint.

### 2.3. Data sources and measurement

According to the internal rules of the clinic, all paper-based patient records were disinfected after discharge and stored in a specially ventilated room; all disinfected records were included. The records were accessed for research purposes between 15 September and 30 October 2020. Epidemiological characteristics, clinical data, laboratory results and treatment during hospitalization were retrieved from the records by two trained researchers and coded by the author with double-checking by a data scientist. All records were fully de-identified before analysis, and the authors did not have access to information that could identify individual participants during or after data collection.

### 2.4. Statistical analysis

Categorical variables are presented as frequencies with percentages and were compared using the chi-square test or Fisher’s exact test (when the expected cell count was < 5 in more than one-fifth of cells). Continuous variables are presented as the median with interquartile range (IQR); normally distributed variables were compared using the Student’s t-test and otherwise using the Mann-Whitney U test. The association of candidate covariates - including sex, age, comorbidities, complications and symptoms - with ICU admission was first examined using univariable binary logistic regression. A multivariable logistic regression model was then fitted, entering clinically pre-specified predictors (age, sex, diabetes mellitus, ischemic heart disease, hypertensive heart disease, pneumonia, shortness of breath, white-blood-cell count and haemoglobin) simultaneously; the composite “any comorbidity” and “any complication” terms were excluded from the multivariable model to avoid collinearity with their components. Crude and adjusted odds ratios (OR and aOR) with 95% confidence intervals (CI) are reported, and model discrimination was summarized by the area under the receiver-operating-characteristic curve (AUC). A P-value < 0.05 was considered statistically significant. Analyses were performed in SPSS version 26 (IBM SPSS Inc., Armonk, NY, USA).

### 2.5. Ethics

The Review Board of the Ministry of Health of the Republic of Uzbekistan reviewed and approved the protocol for this study (No. 6/1-1444, 30 October 2020). Because the study used de-identified records collected during routine care, the requirement for individual informed consent was waived.

## 3. Results

Between 11 April and 8 August 2020, 2500 patients with COVID-19 were admitted to the Multidisciplinary Clinic of the Tashkent State Medical University (Fig. 1). Of these, 989 (39.6%) were asymptomatic and 1511 (60.4%) had at least one symptom. The median age was 36 years (IQR 26-51; range 1-92); 978 patients (39.1%) were women, including 31 (1.2%) who were pregnant. The median time from symptom onset to admission was 7 days (IQR 5-10). The median incubation period differed between groups (8 days [IQR 6-8] in the asymptomatic group and 5 days [IQR 2-7] in the symptomatic group). Baseline characteristics and their distribution by symptom status are shown in Table 1 and Fig. 2.

**Figure 1.**
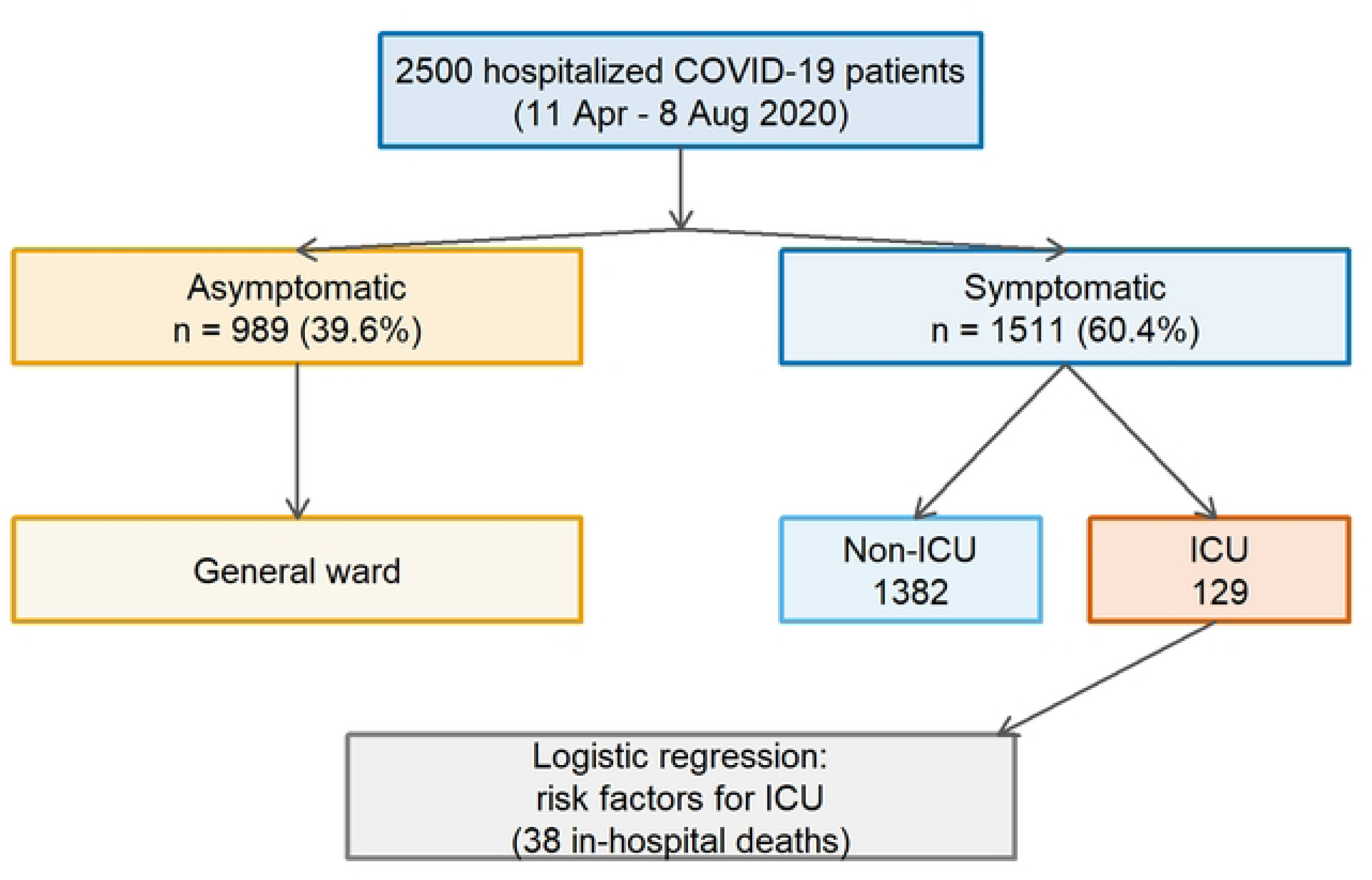
Patient flow and analytic groups. Of 2500 hospitalized COVID-19 patients, 989 (39.6%) were asymptomatic and 1511 (60.4%) symptomatic; 129 symptomatic patients (8.5%) were admitted to the intensive care unit (ICU) and 38 died in hospital.

**Figure 2.**
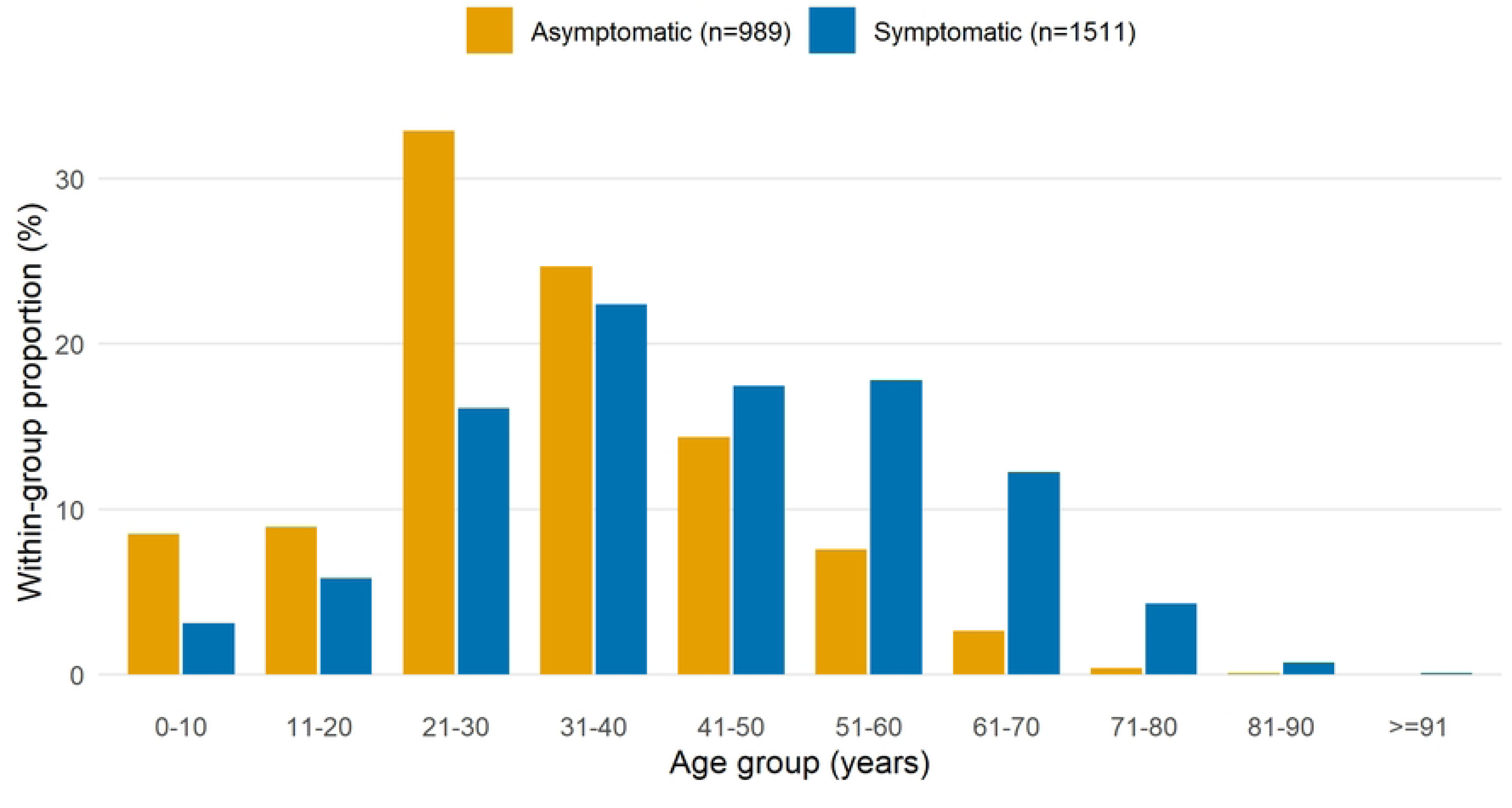
Within-group age distribution of asymptomatic and symptomatic patients. Asymptomatic infections were concentrated in younger adults, whereas symptomatic patients were older.

**Table 1.** Baseline characteristics of 2500 COVID-19 patients.

| Characteristic | Total (n=2500) | Asymptomatic (n=989) | Symptomatic (n=1511) |
| --- | --- | --- | --- |
| Age, median (IQR) | 36 (26-51) | 30 (22-41) | 42 (30-56) |
| Age group, years, n (%) |  |  |  |
| 0-10 | 131 (5.2) | 84 (8.5) | 47 (3.1) |
| 11-20 | 176 (7.0) | 88 (8.9) | 88 (5.8) |
| 21-30 | 568 (22.7) | 325 (32.9) | 243 (16.1) |
| 31-40 | 582 (23.3) | 244 (24.7) | 338 (22.4) |
| 41-50 | 406 (16.2) | 142 (14.4) | 264 (17.5) |
| 51-60 | 344 (13.8) | 75 (7.6) | 269 (17.8) |
| 61-70 | 211 (8.4) | 26 (2.6) | 185 (12.2) |
| 71-80 | 69 (2.8) | 4 (0.4) | 65 (4.3) |
| 81-90 | 12 (0.5) | 1 (0.1) | 11 (0.7) |
| ≥91 | 1 (0.04) | - | 1 (0.1) |
| Sex, n (%) |  |  |  |
| male | 1522 (60.9) | 729 (73.7) | 793 (52.5) |
| female | 978 (39.1) | 260 (26.3) | 718 (47.5) |
| pregnant | 31 (1.2) | 21 (2.1) | 10 (0.7) |
| Treatment place, n (%) |  |  |  |
| general ward | 2370 (94.8) | 989 (100) | 1382 (91.5) |
| ICU | 129 (5.2) | - | 129 (8.5) |
| History of exposure, n (%) |  |  |  |
| from abroad | 703 (28.1) | 478 (48.3) | 225 (14.9) |
| direct contact with a patient | 356 (14.2) | 83 (8.4) | 273 (18.1) |
| family cluster | 396 (15.8) | 150 (15.2) | 246 (16.3) |
| unknown | 776 (31.0) | 117 (11.8) | 659 (43.6) |
| contact with a suspected person | 269 (10.8) | 161 (16.3) | 108 (7.1) |
| Symptoms, n (%) |  |  |  |
| none | 989 (39.6) | 989 (100) | - |
| 1 | 205 (8.2) | - | 205 (13.6) |
| 2 | 261 (10.4) | - | 261 (17.3) |
| 3 or more | 1039 (41.6) | - | 1039 (68.8) |
| severe, cannot describe | 6 (0.2) | - | 6 (0.4) |
| Respiratory rate, /min, median (IQR) | 19 (18-21) | 18 (18-20) | 20 (18-23) |
| Oxygen saturation, %, median (IQR) | 97 (94-98) | 97 (96-98) | 96 (92-98) |
| Comorbidities, n (%) |  |  |  |
| none | 2124 (85.0) | 970 (98.1) | 1154 (76.4) |
| 1 | 175 (7.0) | 13 (1.3) | 162 (10.7) |
| 2 | 60 (2.4) | 3 (0.3) | 57 (3.8) |
| 3 or more | 141 (5.6) | 3 (0.3) | 138 (9.1) |
| Treatment, n (%) |  |  |  |
| antiviral | 2024 (81.0) | 945 (95.6) | 1079 (71.4) |
| antibiotic | 2432 (97.3) | 965 (97.6) | 1467 (97.1) |
| anticoagulant | 786 (31.4) | 19 (1.9) | 767 (50.8) |
| mucolytic | 908 (36.3) | 86 (8.7) | 822 (54.4) |
| antifungal | 214 (8.6) | 48 (4.9) | 166 (11.0) |
| hormonal | 842 (33.7) | 30 (3.0) | 812 (53.7) |
| vitamin | 2324 (93.0) | 937 (94.7) | 1387 (91.8) |
| Outcome, n (%) |  |  |  |
| discharged to home isolation | 1583 (63.3) | 518 (52.4) | 1065 (70.5) |
| transferred to rehabilitation | 833 (33.3) | 470 (47.5) | 363 (24.0) |
| transferred to another hospital | 34 (1.4) | 1 (0.1) | 33 (2.2) |
| deceased | 38 (1.5) | - | 38 (2.5) |
| refused treatment | 12 (0.5) | - | 12 (0.8) |
| Onset-to-admission, days, median (IQR) | 7 (5-10) | 6 (4-10) | 7 (5-10) |
| Incubation period, days, median (IQR) | 5 (2-7.5) | 8 (6-8) | 5 (2-7) |

*IQR, interquartile range; ICU, intensive care unit.*

The symptomatic group was divided into non-ICU and ICU patients. Overall, 129 patients (5.2% of all admissions; 8.5% of symptomatic patients) were admitted to the ICU (Table 2). ICU patients were older (median 56 [IQR 44.5-65] vs 40.5 [IQR 30-55] years). The clinical classification differed markedly between groups: mild (49.6%) and moderate (47.9%) illness predominated among non-ICU patients, whereas severe (34.9%) and critical (37.2%) illness dominated the ICU group. The most common symptoms among symptomatic patients were fatigue (83.2%), cough (58.5%), fever (56.6%), shortness of breath (39.9%) and headache (16.5%). ICU patients more frequently reported fatigue, shortness of breath, cough and loss of appetite (Fig. 3). A respiratory rate of 25-30 breaths/min was recorded in 67 (51.9%) ICU patients, and 61 (47.3%) had oxygen saturation below 79%. Comorbidities including hypertensive heart disease (41.1%), stable angina (39.5%), hypertension (35.6%) and diabetes mellitus (21.7%) were more common in the ICU group.

**Figure 3.**
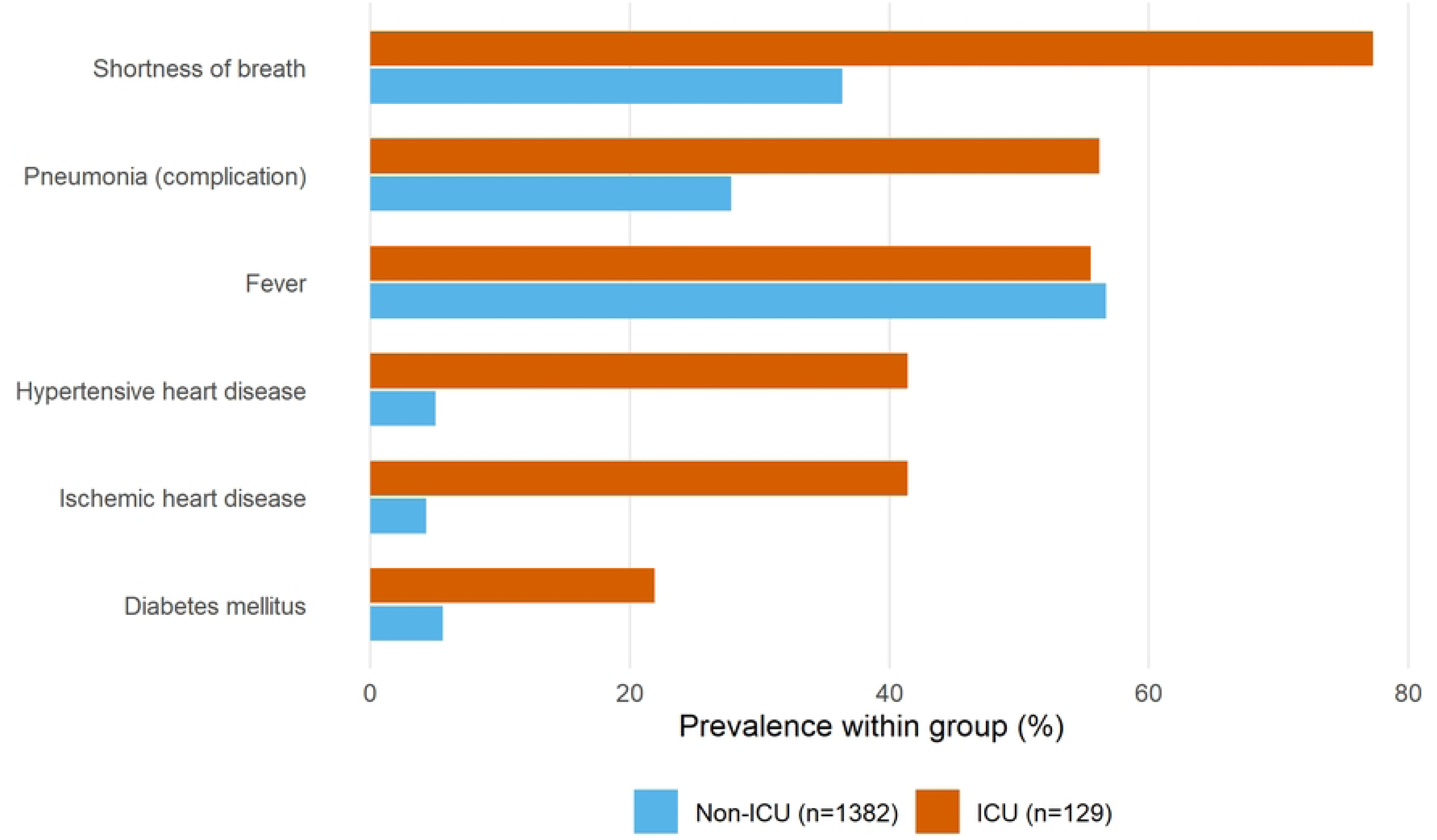
Prevalence of selected symptoms, radiographic findings and comorbidities in ICU versus non-ICU symptomatic patients. All comparisons P < 0.01 except cough and fatigue (P < 0.005).

**Table 2.** Clinical characteristics of symptomatic patients by ICU status.

| Characteristic | Symptomatic (n=1511) | Non-ICU (n=1382) | ICU (n=129) | P |
| --- | --- | --- | --- | --- |
| Age, median (IQR) | 42 (30-56) | 40.5 (30-55) | 56 (44.5-65) | <0.001 |
| Age group, n (%) |  |  |  | <0.001 |
| ≤17 | 73 (4.8) | 73 (5.3) | - |  |
| 18-40 | 643 (42.6) | 618 (44.7) | 25 (19.4) |  |
| 41-65 | 639 (42.3) | 566 (41.0) | 73 (56.6) |  |
| ≥66 | 156 (10.3) | 125 (9.0) | 31 (24.0) |  |
| Sex, n (%) |  |  |  | 0.178 |
| male | 793 (52.5) | 718 (52.0) | 75 (58.1) |  |
| female | 718 (47.5) | 664 (48.0) | 54 (41.9) |  |
| Clinical classification, n (%) |  |  |  | <0.001 |
| mild | 688 (45.5) | 685 (49.6) | 3 (2.3) |  |
| moderate | 695 (46.0) | 662 (47.9) | 33 (25.6) |  |
| severe | 73 (4.8) | 28 (2.0) | 45 (34.9) |  |
| critical | 55 (3.6) | 7 (0.5) | 48 (37.2) |  |
| Selected symptoms, n (%) |  |  |  |  |
| fever | 855 (56.6) | 783 (56.7) | 72 (55.8) | 0.853 |
| fatigue | 1257 (83.2) | 1138 (82.3) | 119 (92.2) | 0.004 |
| headache | 250 (16.5) | 212 (15.3) | 38 (29.5) | <0.001 |
| shortness of breath | 603 (39.9) | 504 (36.5) | 99 (76.7) | <0.001 |
| cough | 884 (58.5) | 788 (57.0) | 96 (74.4) | <0.001 |
| loss of appetite | 143 (9.5) | 97 (7.0) | 46 (35.7) | <0.001 |
| chest pain | 99 (6.6) | 82 (5.9) | 17 (13.2) | 0.001 |
| tachycardia | 42 (2.8) | 25 (1.8) | 17 (13.2) | <0.001 |
| Respiratory rate, n (%) |  |  |  | <0.001 |
| <20 | 877 (58.0) | 863 (62.4) | 14 (10.9) |  |
| 21-24 | 419 (27.7) | 385 (27.9) | 34 (26.4) |  |
| 25-30 | 195 (12.9) | 128 (9.3) | 67 (51.9) |  |
| ≥31 | 20 (1.3) | 6 (0.4) | 14 (10.9) |  |
| Oxygen saturation, n (%) |  |  |  | <0.001 |
| >95 | 911 (60.3) | 894 (64.7) | 17 (13.2) |  |
| 90-94 | 401 (26.5) | 374 (27.1) | 27 (20.9) |  |
| 85-89 | 85 (5.6) | 72 (5.2) | 13 (10.1) |  |
| 80-84 | 33 (2.2) | 22 (1.6) | 11 (8.5) |  |
| <79 | 81 (5.4) | 20 (1.4) | 61 (47.3) |  |
| Comorbidities, n (%) |  |  |  |  |
| stable angina | 102 (6.7) | 51 (4.0) | 51 (39.5) | <0.001 |
| diabetes mellitus | 106 (7.0) | 78 (5.6) | 28 (21.7) | <0.001 |
| hypertensive heart disease | 122 (8.0) | 69 (5.0) | 53 (41.1) | <0.001 |
| hypertension | 102 (6.7) | 56 (4.0) | 46 (35.6) | <0.001 |
| obesity | 28 (1.8) | 4 (0.3) | 24 (18.6) | <0.001 |
| X-ray, n (%) |  |  |  | <0.001 |
| negative | 169 (11.2) | 167 (12.1) | 2 (1.6) |  |
| bronchitis | 393 (26.0) | 382 (27.6) | 11 (8.5) |  |
| bilateral pneumonia | 446 (29.5) | 364 (26.3) | 82 (63.6) |  |
| bilateral bronchopneumonia | 280 (18.5) | 260 (18.8) | 20 (15.5) |  |
| Laboratory, median (IQR) |  |  |  |  |
| WBC, 10<sup>9</sup>/L | 6.3 (4.9-7.8) | 6.2 (4.9-7.7) | 7.1 (5.5-8.9) | <0.001 |
| lymphocytes, % | 27.2 (20.3-33.5) | 28 (21.7-34.1) | 20 (14.5-25.8) | <0.001 |
| ESR, mm/h | 10 (5-17) | 10 (7-17) | 18 (10-25) | <0.001 |
| creatinine, µmol/L | 74.4 (65.9-84.6) | 73.2 (65.4-84) | 80.1 (70.5-105.8) | <0.001 |
| ALT, U/L | 34.2 (27.3-44.1) | 34.1 (27.3-42.4) | 38.9 (29.2-60.1) | <0.001 |
| AST, U/L | 27 (16-33) | 26 (16-32.5) | 31 (20-38.7) | 0.003 |
| fibrinogen, mg/dL | 301 (244-375) | 310 (266-377) | 333 (288-399.7) | 0.007 |
| Treatment, n (%) |  |  |  | <0.001 |
| antiviral | 1079 (71.4) | 1004 (72.6) | 75 (58.1) |  |
| antibiotic | 1467 (97.1) | 1354 (98.0) | 113 (87.6) |  |
| anticoagulant | 767 (50.8) | 662 (47.9) | 105 (81.4) |  |
| hormonal | 812 (53.7) | 722 (52.2) | 90 (69.8) |  |
| Selected complications, n (%) |  |  |  |  |
| bilateral pneumonia | 422 (27.9) | 355 (25.7) | 67 (51.9) | <0.001 |
| type II respiratory failure | 165 (10.9) | 112 (8.1) | 53 (41.1) | <0.001 |
| type III respiratory failure | 55 (3.6) | 9 (0.7) | 46 (35.7) | <0.001 |
| acute heart failure | 50 (3.3) | 24 (1.7) | 26 (20.2) | <0.001 |
| ARDS | 8 (0.5) | 1 (0.1) | 7 (5.4) | <0.001 |
| Outcome, n (%) |  |  |  | <0.001 |
| discharged to home isolation | 1065 (70.5) | 1019 (73.7) | 46 (35.7) |  |
| transferred to rehabilitation | 363 (24.0) | 338 (24.5) | 25 (19.4) |  |
| deceased | 38 (2.5) | - | 38 (29.5) |  |
| Length of stay, days, median (IQR) | 10 (8-12) | 10 (8-12) | 11 (6-17) | 0.019 |

*IQR, interquartile range; ICU, intensive care unit; WBC, white blood cells; ESR, erythrocyte sedimentation rate; ALT, alanine aminotransferase; AST, aspartate aminotransferase; ARDS, acute respiratory distress syndrome. P-values from the chi-square/Fisher's exact test (categorical) or Mann-Whitney U test (continuous).*

Bilateral pneumonia on X-ray was present in 82 (63.6%) ICU patients. Compared with the non-ICU group, the ICU group had higher white-blood-cell counts, platelets, erythrocyte sedimentation rate, creatinine, alanine and aspartate aminotransferase and fibrinogen (Table 2). The most prescribed treatments among symptomatic patients were antibiotics (97.1%), vitamins (91.8%), antivirals (71.4%), mucolytics (54.4%), hormonal therapy (53.7%) and anticoagulants (50.8%); ICU patients more often received antibiotics, anticoagulants and hormonal therapy.

In univariable logistic regression, higher odds of ICU admission were associated with older age (OR 1.048, 95% CI 1.036-1.060), any comorbidity (OR 15.095, 9.770-23.321), ischemic heart disease (OR 15.638, 10.101-24.210), hypertensive heart disease (OR 13.270, 8.665-20.322), diabetes mellitus (OR 4.635, 2.877-7.467), complications (OR 9.978, 5.672-17.553), pneumonia (OR 3.271, 2.267-4.721), shortness of breath (OR 5.749, 3.766-8.775) and white-blood-cell count (OR 1.147, 1.083-1.214), whereas haemoglobin was inversely associated (OR 0.987, 0.977-0.997). Male sex, chronic obstructive pulmonary disease, fever and time from symptom onset to admission were not significantly associated with ICU admission (Table 3, Fig. 4).

**Figure 4.**
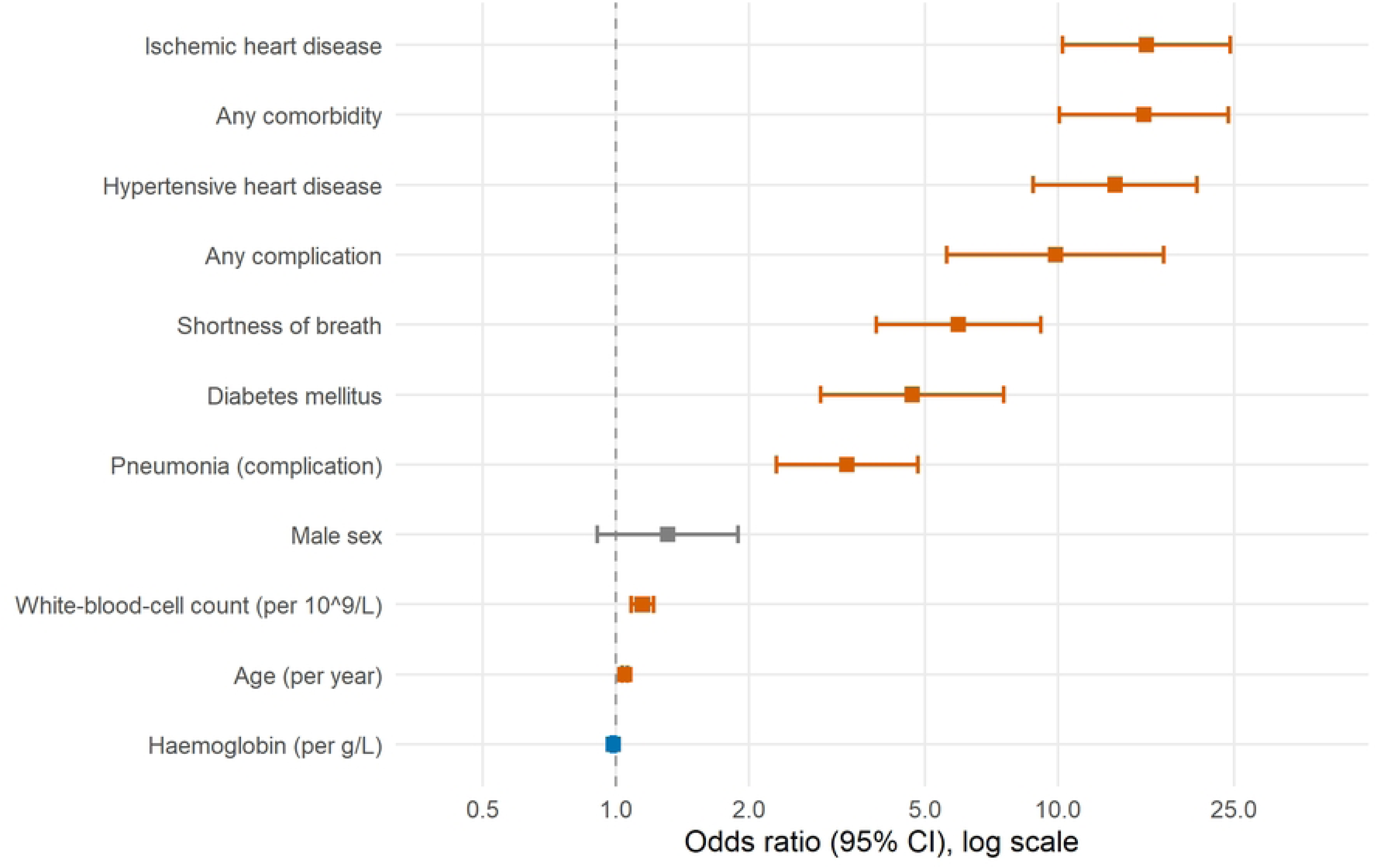
Crude (univariable) odds ratios (95% confidence intervals) for ICU admission (Table 3), plotted on a logarithmic scale. Estimates to the right of the reference line (OR = 1) indicate higher odds of ICU admission; adjusted estimates from the multivariable model are given in Table 3.

**Table 3.** Factors associated with ICU admission: univariable and multivariable logistic regression.

| Covariate | Crude OR (95% CI) | P | Adjusted OR (95% CI) | P |
| --- | --- | --- | --- | --- |
| Male sex | 1.284 (0.891-1.851) | 0.179 | 2.003 (1.243-3.229) | 0.004 |
| Age (per year) | 1.048 (1.036-1.060) | <0.001 | 1.005 (0.990-1.020) | 0.538 |
| Any comorbidity | 15.095 (9.770-23.321) | <0.001 | - | - |
| Ischemic heart disease | 15.638 (10.101-24.210) | <0.001 | 4.199 (2.154-8.185) | <0.001 |
| Diabetes mellitus | 4.635 (2.877-7.467) | <0.001 | 1.586 (0.859-2.929) | 0.141 |
| Hypertensive heart disease | 13.270 (8.665-20.322) | <0.001 | 2.933 (1.507-5.708) | 0.002 |
| Chronic obstructive pulmonary disease | 4.337 (0.833-22.580) | 0.081 | - | - |
| Any complication | 9.978 (5.672-17.553) | <0.001 | - | - |
| Pneumonia | 3.271 (2.267-4.721) | <0.001 | 1.168 (0.716-1.904) | 0.533 |
| Onset-to-admission (per day) | 0.972 (0.925-1.021) | 0.260 | - | - |
| Fever | 0.966 (0.672-1.390) | 0.853 | - | - |
| Shortness of breath | 5.749 (3.766-8.775) | <0.001 | 3.224 (1.878-5.535) | <0.001 |
| White-blood-cell count (per 10<sup>9</sup>/L) | 1.147 (1.083-1.214) | <0.001 | 1.038 (0.965-1.117) | 0.320 |
| Haemoglobin (per g/L) | 0.987 (0.977-0.997) | 0.009 | 0.999 (0.986-1.013) | 0.929 |

OR, odds ratio; CI, confidence interval; ICU, intensive care unit. Crude estimates are from univariable models. Adjusted estimates are from a single multivariable model (n = 1470; 121 ICU events; AUC 0.82) entering age, sex, diabetes mellitus, ischemic heart disease, hypertensive heart disease, pneumonia, shortness of breath, white-blood-cell count and haemoglobin simultaneously; composite comorbidity/complication terms and variables with substantial missingness (onset-to-admission) were not entered (-).

In the multivariable model (n = 1470 with complete data, 121 ICU events; AUC 0.82), four factors remained independently associated with ICU admission: ischemic heart disease (aOR 4.20, 95% CI 2.15-8.19), shortness of breath (aOR 3.22, 1.88-5.54), hypertensive heart disease (aOR 2.93, 1.51-5.71) and male sex (aOR 2.00, 1.24-3.23). After mutual adjustment, the associations of age (aOR 1.01, 0.99-1.02), diabetes mellitus (aOR 1.59, 0.86-2.93), pneumonia (aOR 1.17, 0.72-1.90), white-blood-cell count and haemoglobin were attenuated and no longer significant, indicating that much of their crude association was explained by co-occurring cardiac comorbidity and respiratory compromise (Table 3).

## 4. Discussion

In this cross-sectional study of 2500 hospitalized COVID-19 patients from the first pandemic wave in Uzbekistan, we described the clinical spectrum of the disease and identified risk factors for ICU admission. Older age, cardiometabolic comorbidity, respiratory compromise and elevated white-cell count were the principal correlates of intensive care. To our knowledge, this is one of the first detailed clinical COVID-19 datasets reported from Uzbekistan and Central Asia, a region that remains largely absent from the international literature; the description therefore has value as a regional baseline even though the data derive from 2020, because it captures the clinical presentation and hospital response of a resource-constrained system before disease-specific therapies and vaccines were available.

Of the 2500 patients, 1522 (60.9%) were men and 978 (39.1%) were women, including 31 (1.2%) pregnant women. Similar to Breslin et al.,^11^ most pregnant women were asymptomatic and none were admitted to the ICU. Consistent with previous studies,^12,13^ men predominated in both the asymptomatic (73.7%) and symptomatic (52.5%) groups. A total of 989 (39.6%) patients were laboratory-confirmed asymptomatic infections; as others have noted,^14,15^ distinguishing asymptomatic from presymptomatic patients requires chronological observation, and in our study the absence of symptoms persisted throughout hospitalization.

Most asymptomatic patients (48.3%) were infected abroad, whereas this applied to only 14.9% of symptomatic patients; the route of transmission was unknown for 43.6% of symptomatic patients, probably because early efforts focused on isolating symptomatic cases while presymptomatic carriers escaped attention.^20,21^ Family transmission was observed in 16.3% of symptomatic and 15.2% of asymptomatic patients, in keeping with reports that early home isolation, presymptomatic transmission and close contact contribute to family clusters.^22,23,24^

When epidemiological and clinical features were compared between non-ICU and ICU patients, clinical manifestations were more pronounced in the ICU group. Based on the nationwide Korean database, Park et al. similarly reported that symptomatic patients were more likely to require intensive care than asymptomatic patients.^27^ Comorbidities such as hypertensive heart disease, stable angina, hypertension and diabetes mellitus were most common in the ICU group,^17,29^ and bilateral pulmonary involvement predominated on imaging, consistent with Huang et al.^3^ Laboratory analysis showed higher erythrocyte sedimentation rate and aminotransferase levels in the ICU group, in agreement with Yang et al.^30^ and Zhou et al.^31^

At the beginning of the pandemic no specific therapy was available.^32^ Antibiotic, vitamin and antiviral therapy were given to the majority of non-ICU patients according to the interim guidelines, while hormonal therapy was more frequent in the ICU group owing to disease severity. Complications were also more frequent in the ICU group, including bilateral pneumonia (51.9%), type II (41.1%) and type III (35.7%) respiratory failure, acute heart failure (20.2%) and acute respiratory distress syndrome (5.4%), consistent with previous reports.^1,3,33^ The median hospital stay was 10 days (IQR 8-12) in the non-ICU group and 11 days (IQR 6-17) in the ICU group; 38 ICU patients (29.5%) died.^34^

Binary logistic regression indicated that male sex, chronic obstructive pulmonary disease, fever and time from symptom onset to admission were not significantly associated with ICU admission, in line with several previous reports.^35,36,37^ Consistent with other studies,^38,39^ older age, comorbidities (ischemic heart disease, diabetes mellitus and hypertensive heart disease), complications, pneumonia, shortness of breath and changes in white-cell count and haemoglobin increased the probability of ICU admission.

### 4.1. Limitations

This study has several limitations. First, it was conducted at a single centre, so the risk-factor estimates are most representative of that hospital. Second, the cross-sectional design and the reliance on admission-time variables limit causal and temporal inference. Third, the data derive from the first pandemic wave in 2020, before vaccination and before SARS-CoV-2 variants of concern, so the clinical spectrum may differ from that of later waves. Fourth, the number of ICU patients and deaths was limited (129 and 38, respectively); nevertheless, these remain a valuable early description of severe COVID-19 in Uzbekistan. Fifth, although we fitted a multivariable model with pre-specified predictors and excluded collinear composite terms, the number of ICU events (121 with complete data) limits the number of covariates that can be adjusted for simultaneously, so the adjusted estimates should be regarded as exploratory. Finally, some laboratory parameters (for example cytokines and detailed coagulation markers) were not available in the records.

## 5. Conclusion

Among 2500 hospitalized COVID-19 patients during the first wave in Uzbekistan, older age, cardiometabolic comorbidity, pneumonia, shortness of breath and elevated white-cell count were associated with ICU admission, whereas male sex, COPD, fever and time to admission were not. Particular attention should be paid to cases with an unknown route of transmission and to family clusters, and to older patients and those with heart disease or diabetes mellitus, who are at high probability of requiring intensive care. As one of the first clinical COVID-19 descriptions from Central Asia, these findings provide a baseline for the rapid assessment of similar outbreaks and for preventive-health-policy development in the region.

## Data Availability

All relevant aggregated data are within the manuscript and its supporting information files. Patient-level data cannot be shared publicly because they contain potentially identifying clinical information de-identified data are available from the Tashkent State Medical University ethics board (contact: via the corresponding author) for researchers who meet the criteria for access to confidential data.

## Ethical approval

The Review Board of the Ministry of Health of the Republic of Uzbekistan reviewed and approved the study protocol (No. 6/1-1444, 30 October 2020). The requirement for individual informed consent was waived because de-identified routine-care records were used.

## Author contributions

Conceptualization: J.W.C., K.K., B.R. Data curation: L.N.T., A.K.S., B.R. Formal analysis: B.R. Investigation: B.R. Methodology: J.W.C., K.K., B.R. Validation: L.N.T., A.K.S., B.M. Writing – original draft: B.R. Writing – review & editing: B.M., L.N.T., A.K.S. All authors reviewed and approved the final manuscript.

## Data availability

The data that support the findings of this study are available from the corresponding author upon reasonable request. They are not publicly available because they contain information that could compromise the privacy of research participants.

## Declaration of generative AI and AI-assisted technologies

During the preparation of this work the authors used AI-assisted tools in order to improve the language and readability of the manuscript. After using these tools, the authors reviewed and edited the content as needed and take full responsibility for the content of the publication.

## Acknowledgements

The authors thank the Tashkent State Medical University and its Multidisciplinary Clinic for their contribution to data collection.

## References

1. Chen NS, Zhou M, Dong X, Qu JM, Gong FY, Han Y, et al. Epidemiological and clinical characteristics of 99 cases of 2019 novel coronavirus pneumonia in Wuhan, China: a descriptive study. Lancet 2020;395(10223):507–13.

2. WHO Director-General’s remarks at the media briefing on 2019-nCoV on 11 February 2020. Available from: https://www.who.int/director-general/speeches/detail/who-director-general-s-remarks-at-the-media-briefing-on-2019-ncov-on-11-february-2020. Accessed 20 January 2022.

3. Huang C, Wang Y, Li X, Ren L, Zhao J, Hu Y, et al. Clinical features of patients infected with 2019 novel coronavirus in Wuhan, China. Lancet 2020;395(10223):497–506.

4. Teich VD, Klajner S, Almeida FAS, Dantas ACB, Laselva CR, Torritesi MG, et al. Epidemiologic and clinical features of patients with COVID-19 in Brazil. Einstein (Sao Paulo) 2020;18:eAO6022.

5. Immovilli P, Morelli N, Antonucci E, Radaelli G, Barbera M, Guidetti D. COVID-19 mortality and ICU admission: the Italian experience. Crit Care 2020;24(1):228.

6. Jain V, Yuan JM. Predictive symptoms and comorbidities for severe COVID-19 and intensive care unit admission: a systematic review and meta-analysis. Int J Public Health 2020;65(5):533–46.

7. OECD. Strengthening the frontline: How primary health care helps health systems adapt during the COVID-19 pandemic. Paris: OECD; 2021. Accessed 21 January 2022.

8. Kim K, Choi JW, Moon J, Akilov H, Tuychiev L, Rakhimov B, et al. Clinical Features of COVID-19 in Uzbekistan. J Korean Med Sci 2020;35(45):e404.

9. Interim guidelines for the management of patients infected with COVID-19 (6th ed.). The Ministry of Health of the Republic of Uzbekistan; 2020.

10. World Health Organization. Responding to the COVID-19 pandemic: WHO’s action in countries, territories and areas, 2020. Geneva: WHO; 2021. Licence: CC BY-NC-SA 3.0 IGO.

11. Breslin N, Baptiste C, Gyamfi-Bannerman C, Miller R, Martinez R, Bernstein K, et al. Coronavirus disease 2019 infection among asymptomatic and symptomatic pregnant women: two weeks of confirmed presentations to an affiliated pair of New York City hospitals. Am J Obstet Gynecol MFM 2020;2(2):100118.

12. Shahriarirad R, Khodamoradi Z, Erfani A, Hosseinpour H, Ranjbar K, Emami Y, et al. Epidemiological and clinical features of 2019 novel coronavirus diseases (COVID-19) in the South of Iran. BMC Infect Dis 2020;20(1):427.

13. Wu B, Lei ZY, Wu KL, He JR, Cao HJ, Fu J, et al. Compare the epidemiological and clinical features of imported and local COVID-19 cases in Hainan, China. Infect Dis Poverty 2020;9(1):143.

14. Workman J. The proportion of COVID-19 cases that are asymptomatic in South Korea: Comment on Nishiura et al. Int J Infect Dis 2020;96:398.

15. Jung CY, Park H, Kim DW, Choi YJ, Kim SW, Chang TI. Clinical Characteristics of Asymptomatic Patients with COVID-19: A Nationwide Cohort Study in South Korea. Int J Infect Dis 2020;99:266–8.

16. Grasselli G, Zangrillo A, Zanella A, Antonelli M, Cabrini L, Castelli A, et al. Baseline Characteristics and Outcomes of 1591 Patients Infected With SARS-CoV-2 Admitted to ICUs of the Lombardy Region, Italy. JAMA 2020;323(16):1574–81.

17. Guan WJ, Ni ZY, Hu Y, Liang WH, Ou CQ, He JX, et al. Clinical Characteristics of Coronavirus Disease 2019 in China. N Engl J Med 2020;382(18):1708–20.

18. Stokes EK, Zambrano LD, Anderson KN, Marder EP, Raz KM, El Burai Felix S, et al. Coronavirus Disease 2019 Case Surveillance - United States, January 22-May 30, 2020. MMWR Morb Mortal Wkly Rep 2020;69(24):759–65.

19. Zhao Z, Chen A, Hou W, Graham JM, Li H, Richman PS, et al. Prediction model and risk scores of ICU admission and mortality in COVID-19. PLoS One 2020;15(7):e0236618.

20. Tindale LC, Stockdale JE, Coombe M, Garlock ES, Lau WYV, Saraswat M, et al. Evidence for transmission of COVID-19 prior to symptom onset. Elife 2020;9:e57149.

21. He X, Lau EHY, Wu P, Deng X, Wang J, Hao X, et al. Temporal dynamics in viral shedding and transmissibility of COVID-19. Nat Med 2020;26(5):672–5.

22. Qiu YY, Wang SQ, Wang XL, Lu WX, Qiao D, Li JB, et al. Epidemiological analysis on a family cluster of COVID-19. Zhonghua Liu Xing Bing Xue Za Zhi 2020;41(4):494–7.

23. Qian G, Yang N, Ma AHY, Wang L, Li G, Chen X, et al. COVID-19 Transmission Within a Family Cluster by Presymptomatic Carriers in China. Clin Infect Dis 2020;71(15):861–2.

24. Yong SEF, Anderson DE, Wei WE, Pang J, Chia WN, Tan CW, et al. Connecting clusters of COVID- 19: an epidemiological and serological investigation. Lancet Infect Dis 2020;20(7):809–15.

25. Chen J, Qi T, Liu L, Ling Y, Qian Z, Li T, et al. Clinical progression of patients with COVID-19 in Shanghai, China. J Infect 2020;80(5):e1–e6.

26. Xie H, Zhao J, Lian N, Lin S, Xie Q, Zhuo H. Clinical characteristics of non-ICU hospitalized patients with coronavirus disease 2019 and liver injury: A retrospective study. Liver Int 2020;40(6):1321–6.

27. Park HC, Kim DH, Cho A, Kim J, Yun KS, Kim J, et al. Clinical outcomes of initially asymptomatic patients with COVID-19: a Korean nationwide cohort study. Ann Med 2021;53(1):357–64.

28. Wang F, Yang Y, Dong K, Yan Y, Zhang S, Ren H, et al. Clinical Characteristics of 28 Patients with Diabetes and Covid-19 in Wuhan, China. Endocr Pract 2020;26(6):668–74.

29. Wang D, Hu B, Hu C, Zhu F, Liu X, Zhang J, et al. Clinical Characteristics of 138 Hospitalized Patients With 2019 Novel Coronavirus-Infected Pneumonia in Wuhan, China. JAMA 2020;323(11):1061–9.

30. Yang J, Wu K, Ding A, Li L, Lu H, Zhu W, et al. Clinical characteristics, treatment, and prognosis of 74 2019 novel coronavirus disease patients in Hefei: A single-center retrospective study. Medicine (Baltimore) 2021;100(21):e25645.

31. Zhou J, Sun J, Cao Z, Wang W, Huang K, Zheng F, et al. Epidemiological and clinical features of 201 COVID-19 patients in Changsha city, Hunan, China. Medicine (Baltimore) 2020;99(34):e21824.

32. Wiersinga WJ, Rhodes A, Cheng AC, Peacock SJ, Prescott HC. Pathophysiology, Transmission, Diagnosis, and Treatment of Coronavirus Disease 2019 (COVID-19): A Review. JAMA 2020;324(8):782–93.

33. Lei S, Jiang F, Su W, Chen C, Chen J, Mei W, et al. Clinical characteristics and outcomes of patients undergoing surgeries during the incubation period of COVID-19 infection. EClinicalMedicine 2020;21:100331.

34. Rees EM, Nightingale ES, Jafari Y, Waterlow NR, Clifford S, Pearson CAB, et al. COVID-19 length of hospital stay: a systematic review and data synthesis. BMC Med 2020;18(1):270.

35. Yu C, Lei Q, Li W, Wang X, Li W, Liu W. Epidemiological and clinical characteristics of 1663 hospitalized patients infected with COVID-19 in Wuhan, China: a single-center experience. J Infect Public Health 2020;13(9):1202-9.

36. Machado-Alba JE, Valladales-Restrepo LF, Machado-Duque ME, Gaviria-Mendoza A, Sanchez-Ramirez N, Usma-Valencia AF, et al. Factors associated with admission to the intensive care unit and mortality in patients with COVID-19, Colombia. PLoS One 2021;16(11):e0260169.

37. Correa TD, Midega TD, Timenetsky KT, Cordioli RL, Barbas CSV, Silva Junior M, et al. Clinical characteristics and outcomes of COVID-19 patients admitted to the intensive care unit during the first year of the pandemic in Brazil: a single center retrospective cohort study. Einstein (Sao Paulo) 2021;19:eAO6739.

38. Solmaz I, Ozcaylak S, Alakus OF, Kilic J, Kalin BS, Guven M, et al. Risk factors affecting ICU admission in COVID-19 patients; Could air temperature be an effective factor? Int J Clin Pract 2021;75(3):e13803.

39. Alhumaid S, Al Mutair A, Al Alawi Z, Al Salman K, Al Dossary N, Omar A, et al. Clinical features and prognostic factors of intensive and non-intensive 1014 COVID-19 patients: an experience cohort from Alahsa, Saudi Arabia. Eur J Med Res 2021;26(1):47.

